# Multiple Sclerosis Drives Gastric Cancer via Shared Inflammatory Pathways: A Mendelian Randomization Study

**DOI:** 10.1101/2025.09.01.25334880

**Authors:** Hengyi Liao, Xuhao Wang, Yiji Zhang, Zhiya Zhang, Yin Liao

**Author notes:** Contribute equally to the article (Hengyi Liao, Xuhao Wang, Yiji Zhang). Corresponding author, Yin Liao, The Second People’s Hospital of Meishan City, No.177, Section 1, Longtan Avenue, Huairen Street, Renshou County, Meishan 620500, Sichuan, PR China., Zhiya Zhang, Department of Gastroenterology, Beijing Chest Hospital Capital Medical University Beijing Tuberculosis and Thoracic Tumor Research Institute, Beijing 101149, China., E-mail: Zhang.

## Abstract

**Background:** The causal relationship between Multiple Sclerosis (MS) and Gastric Cancer (GC) remains unclear despite reports suggesting that MS, an autoimmune disease, may contribute to the development of various tumors.

**Methods:** Mendelian Randomization (MR) analysis was employed to explore the potential causal relationship between MS and GC. Subsequently, the GEO database was utilized to identify differentially expressed genes (DEGs) that are commonly associated with both MS and GC, thereby revealing the shared molecular mechanisms underlying these two diseases.

**Results:** The MR analysis indicated that MS significantly increased the risk of GC, demonstrating a positive causal effect. However, reverse analysis from GC to MS did not reveal any significant causal relationships. The sensitivity analysis supported the evidence of a positive causal effect of MS on GC. Transcriptomic data analysis identified shared DEGs between MS and GC, particularly those involved in immune regulation, stromal formation, and cell migration, suggesting that these genes operate through similar biological pathways in both diseases.

**Conclusion:** These findings underscore the intricate interplay between autoimmune disorders and gastrointestinal malignancies, offering potential molecular targets for the personalized management of MS and GC prevention.

## Introduction

Multiple sclerosis (MS) is an autoimmune disorder characterized by the immune system erroneously attacking the myelin sheath surrounding nerve fibers^【1】^, leading to impaired neuronal function**^Error! Reference source not found.^**. MS prevalence has increased worldwide since 2013, and females are twice as likely to develop MS as males^【2】^. The precise etiology of MS remains incompletely understood, but it is believed to arise from the complex interplay of genetic and environmental factors^【4】^, including variations in the regulatory regions of immune-related genes^【2】^, itamin D deficiency**^Error! Reference source not found.^**, pecific viral infections such as Epstein-Barr virus^【6】^, nd smoking**^Error! Reference source not found.^**. ndividuals with MS commonly rely on disease-modifying therapies (DMTs) and immunosuppressants to manage symptoms and delay disease progression of the disease^【9】^. However, immunosuppressants may increase the risk of infection and cancer^【10),(11】^, hich should be taken seriously by MS patients in immune-activated or chronically inflammatory states^【12】^.

The incidence and mortality rate of Gastric cancer (GC) rank fifth globally, respectively, with a fourfold higher prevalence in men than in women, making it a common malignant tumor of the gastrointestinal tract**^Error! Reference source not found.^**. key risk factors for GC include Helicobacter pylori infection, unhealthy dietary patterns (e.g., high-salt diets and frequent intake of pickled foods), smoking, and family history of the disease ^【13】^. Progression from chronic infection-inflammation-atrophy-pyrogenesis-heteroproliferative-gastric cancer is a complex, multistep process^【15】^. Both MS and GC are recognized as chronic inflammatory conditions that play a critical role in promoting tumor development through mechanisms such as enhancing cell proliferation, inhibiting cell death, and supporting angiogenesi^(^_s_^16(), 17】^. Both MS and GC are recognized as chronic inflammatory conditions that play a critical role in promoting tumor development through mechanisms such as enhancing cell proliferation, inhibiting cell death, and supporting angiogenesis.

Genome-Wide Association Studies (GWAS) have been widely utilized to investigate causal relationships between various expos. GWAS can identify genetic markers associated with specific diseases by detecting thousands or even millions of single-nucleotide polymorphisms (SNPs)^【18)(19】^. Using GWAS data, Mendelian randomization (MR) analysis can establish causal links between genetic variations and health outcomes, offering more reliable evidence by minimizing confounding factors and reverse causality issues inherent in traditional observational studies^【20】^. By comparing transcriptomic data from patients with different diseases, we can investigate differential gene expression, identify common regulatory networks, and identify key biomarkers^【21】^.

This study aimed to investigate the potential causal relationships between MS and GC. Initially, we explored the shared genetic basis between MS and GC using GWAS data and established a causal relationship through MR analysis. Subsequently, by analyzing transcriptome sequencing data from patients with MS and GC, we aimed to identify differentially expressed genes (DEGs) and the biological pathways involved in these diseases.

## Materials and Methods

### Study Design and Data Source

For this research, we strictly adhered to the STROBE-MR guidelines to ensure transparency and reproducibility**^Error! Reference source not found.^**. Our data were sourced from the openGWAS database (https://gwas.mrcieu.ac.uk/, accessed date: 17^th^, May, 2024) and FinnGen database (https://r10.risteys.finngen.fi/endpoints/C3_STOMACH_EXALLC, accessed date: 17^th^, May, 2024) ^【22】^, publicly available GWAS databases that include extensive case and control data for MS and GC, ensuring the breadth and representativeness of our sample. We utilized two GWAS datasets related to MS in our study. One dataset (GWAS ID: ebi-a-GCST005531) served as the experimental group, including 14,498 cases and 24,091 controls^【24】^; another dataset (GWAS ID: ieu-a-1024) served as the validation group, including 9,722 cases and 17,376 controls ^【25】^. Additionally, we used a GC-related GWAS dataset from the FinnGen database (finn-b-R10_C3_STOMACH_EXALLC) as the main outcome variable for this analysis, which included 1,423 cases and 314,193 controls. These datasets not only comprised a large number of cases but also originated from the same ethnicity but different regions, which helps reduce the risk of bias due to sample overlap in MR analysis. With this rigorous data selection and analysis method, we aimed to enhance the reliability of our research results, thereby more accurately exploring and understanding the potential genetic and molecular connections between MS and GC.

The primary direction of our MR analysis considered MS as the exposure variable and GC as the outcome variable. This framework helped determine whether MS could potentially lead to GC. Conversely, in the reverse MR analysis, GC was considered the exposure variable, with MS as the outcome. Although this was a secondary analysis direction, it was crucial for validating whether a bidirectional causal relationship exists. In all MR analyses, genetic variations closely associated with the exposure variables were selected as IVs. Our carefully chosen IVs met three fundamental assumptions^【26】^: they were strongly associated with the exposure factors, unassociated with potential confounders, and influenced the outcomes solely through the exposure factors and not via other pathways.

Utilizing bioinformatics approaches, we conducted a detailed analysis of the common differential genes between MS and GC, identifying related biological processes and potential molecular regulatory pathways. Further, by constructing protein-protein interaction networks, we identified key interacting genes.

### IVs Selection and Quality Control

To ensure the efficacy and reliability of the IVs, we undertook the following steps for selection and quality control: First, from large-scale association studies, we extracted single nucleotide polymorphisms (SNPs) significantly associated with the exposure factors of MS and GC. In the forward MR analysis, we set a strict P-value threshold (*P*<5E-08) to select SNPs^【27】^, while in the reverse MR analysis, considering the number of significant SNPs available, we adjusted the threshold to *P*<5E-05. To eliminate the effects of linkage disequilibrium, we set parameters of r²=0.001 and a distance window of 10,000 kb to exclude related SNPs^【28】^. Additionally, we checked the completeness of data for the IVs, supplementing missing data using relevant data from the 1000 Genomes Project. The F-statistic for each SNP (F=beta²/se²) was also calculated to ensure all selected SNPs had an F-value greater than 10, verifying their efficacy as IVs**^Error! Reference source not found.^**. Moreover, we screened for potential confounders using the LDlink database^【29】^, specifically excluding SNPs that might be significantly related to the outcome in different analysis directions, such as excluding SNPs related to gastric cancer, Helicobacter pylori infection, and gastritis in the forward analysis, and excluding SNPs related to multiple sclerosis and neurological disorders in the reverse analysis. This step ensured that the IVs included in the analysis met the three key assumptions of MR, enhancing the reliability and scientific validity of our research results.

### MR and Meta-Analysis

In this study, we primarily used the inverse-variance weighted (IVW) method to perform MR analysis, including both forward and reverse analyses ^【31】^. To comprehensively evaluate the robustness of the MR analysis results, we applied a variety of methods, including MR Egger^【32】^, eighted median^【33】^, simple mode**^Error! Reference source not found.^**, and weighted mode^【34】^. When interpreting the results of the MR analysis, we used two criteria to assess their significance: first, the direction of the causal effect estimates obtained by the five methods must be consistent, either all indicating a positive direction (causal effect value greater than 0) or all indicating a negative direction (causal effect value less than 0); secondly, the *P*-value obtained in the IVW method needed to reach statistical significance (*P_IVW_*<0.05) ^【36】^. To ensure the reliability of the research results, we conducted extensive sensitivity analyses. This included MR-Egger regression, whose intercept test was used to assess the presence of directional pleiotropy^【37】^, i.e., whether the IVs influenced the outcome only through the exposure factor. Additionally, we used the MR-PRESSO method to identify and exclude outlier SNPs, which helped correct estimation biases caused by unusual genetic variations. To assess the heterogeneity among the selected genetic IVs, we applied the *Cochrane’s Q* tes^t ( 38】^. Furthermore, through meta-analysis, we integrated MR analysis results from different data sources, which helped provide more comprehensive and precise evidence. All statistical analyses were completed in R software version 4.3.3, using primarily the Two Sample MR^【39】^and MR-PRESSO packages ^【40】^. We set a P-value of less than 0.05 as the standard for statistically significant differences^【41】^.

### Transcriptome Data Acquisition and Differential Gene Expression Analysis

In this study, we retrieved and downloaded transcriptome data from the GEO database to analyze gene expression changes related to MS and GC. Using “multiple sclerosis” and “gastric cancer” as keywords and specifying the species as human, we selected expression profile analysis data types based on chip technology. We chose the GC patient transcriptome dataset GSE79973 (https://www.ncbi.nlm.nih.gov/geo/query/acc.cgi?acc=GSE79973)^【42】^ and the MS dataset GSE38010 (https://www.ncbi.nlm.nih.gov/geo/query/acc.cgi?acc=GSE38010)^【43】^. By comparing these with healthy control groups corresponding to the diseases, differential expression analysis helped us identify genes with altered expression in these two diseases^【44】^. The results of the differential gene expression analys is were visually displayed through volcano plots and heatmaps, clearly depicting significant changes in gene expression levels^【45】^. Additionally, we obtained the shared differential expressed genes (DEGs) between MS and GC. These shared DEGs provided valuable clues for subsequent studies, potentially involving common pathological mechanisms between the two diseases. To ensure that the selection of differential expressed genes was biologically meaningful, we set selection criteria: a log-fold change (logFC) greater than 1 and a *P*-value less than 0.01. This stringent selection criterion helped ensure that the identified genes were statistically significant and had a sufficient magnitude of change, making them more likely to play a key role in the disease process.

### Enrichment Analysis and Interaction Network Construction of Co-Expressed Differential Genes

A Venn diagram was used to illustrate the shared DEGs between MS and GC. Subsequently, we conducted Gene Ontology (GO)**^Error! Reference source not found.^** and Kyoto Encyclopedia of Genes and Genomes (KEGG) enrichment analyses^【47】^ on these genes using the ClusterProfiler package^【48】^. This analysis covered three aspects: biological processes, cellular components, and molecular functions, with selection criteria set for an adjusted *P*-value less than 0.05. In the results presentation, we particularly emphasized the top 10 results with the lowest P-values in the GO analysis and the top ten pathways in the KEGG pathway analysis. Additionally, we used the STRING database^【49】^ for Protein-Protein Interaction (PPI) analysis and utilized Cytoscape software to visualize the interaction network among genes ^【50】^. By analyzing the degree of connectivity in the network, we identified key interacting genes via the strength of degree in cytoHubba plug-_in_(^51】^.

## Results

In the forward analysis, with MS as the exposure variable, the experimental group (ebi-a-GCST005531) identified 45 IVs closely associated with MS, and the validation group (ieu-a-1024) identified 26(Table S1). In the reverse analysis, with GC as the exposure variable, one related IV was identified in the experimental group directed towards MS, and 5 SNPs were identified in the validation set(Table S2). After excluding confounding factors, all these IVs were used for forward and reverse MR analysis. The bidirectional MR analysis, conducted using the IVW method, demonstrated a statistically significant positive causal relationship between MS (both experimental and validation groups) and GC (Fig. 1A, OR>1, *P_IVW_*<0.05). Since there was only one SNP in the reverse analysis from GC to MS in the experimental group, we used the Wald ratio method to evaluate the causal effect between them, which showed no statistical difference (Fig. S1). Additionally, the reverse MR analysis from GC to the MS validation group also did not show significance (Fig. S1, *P_IVW_*>0.05). Results of MR analysis using five different methods in each direction are listed in Table S3. The meta-analysis results of the forward analysis indicated significance (Fig. 1B, Table S4, *P_meta_*<0.05). These findings emphasize the possibility that MS could influence the occurrence of GC through specific biological pathways, while the impact of GC on MS is weak or not significant, highlighting the importance of causal inference in deciphering interactions between different systemic diseases.

**Fig. 1:**
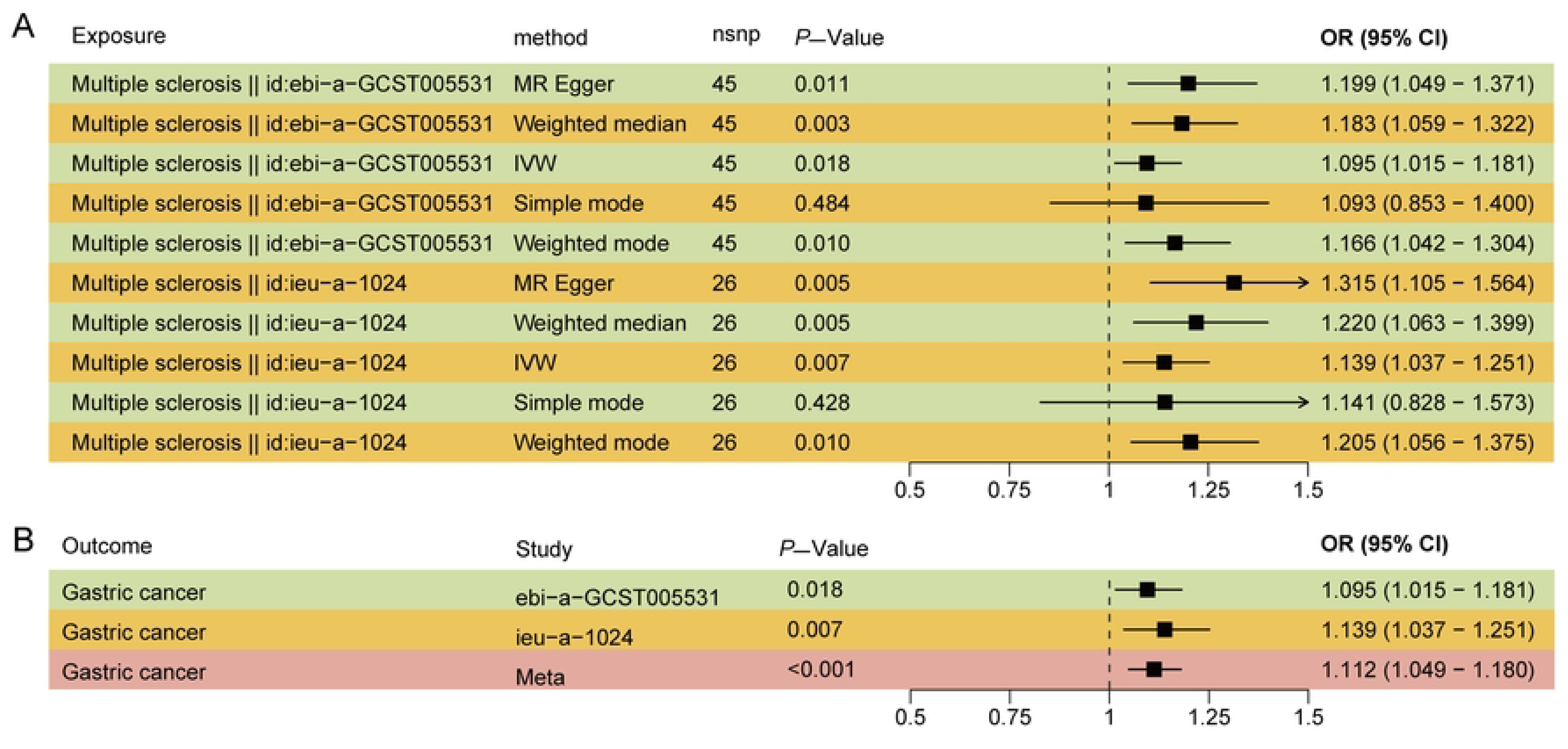
Forest plots show the estimated causal effects (A) of multiple sclerosis (MS) on gastric cancer (GC) and meta-analysis result (B). The position of the black square represents the causal effect values obtained by different MR methods. The short horizontal line passing through the black square represents the 95% confidence interval (CI) of the causal effect value, and the arrow indicates that the upper limit of the CI is outside the range marked in the X axis. nsnp, the number of single nucleotide polymorphisms (SNPs) included in the current analysis; OR, odd ratio; IVW, inverse-variance weighted.

### Sensitivity Analysis

The sensitivity analysis indicated no significant heterogeneity (Table 1, P>0.05) or pleiotropy (Table 1, P>0.05) when MS was considered as the exposure factor and GC as the outcome, for both the experimental and validation groups. Similarly, sensitivity analysis showed no significant heterogeneity (Table 1, *P*>0.05) or pleiotropy (Table 1, *P*>0.05) when GC was the exposure factor and the GC validation group was the outcome. To further assess the impact of individual SNPs included in the analysis on the causal effect estimates, we conducted single SNP effect estimation and leave-one-out analysis. Specifically, in the forward analysis, individual SNPs related to MS in different datasets showed varying trend causal effects on GC (Fig. 2A-B), but the overall effect was significant. This highlights the diversity and complexity of SNPs involved in regulating physiological processes. During the leave-one-out analysis of these SNPs, we found that after removing rs3130283 from the experimental group and rs3134954 from the validation group, the causal effects of the remaining SNPs changed from significant to non-significant, but the overall trend of causal effects remained consistent, positioned to the right of the null effect line (Fig. 2C-D). This indicates that although some SNPs in the current MR analysis exhibit potential bias, the overall results still maintain a certain level of credibility. These findings underscore the complexity of genetic variations in the interactions between diseases.

**Fig. 2:**
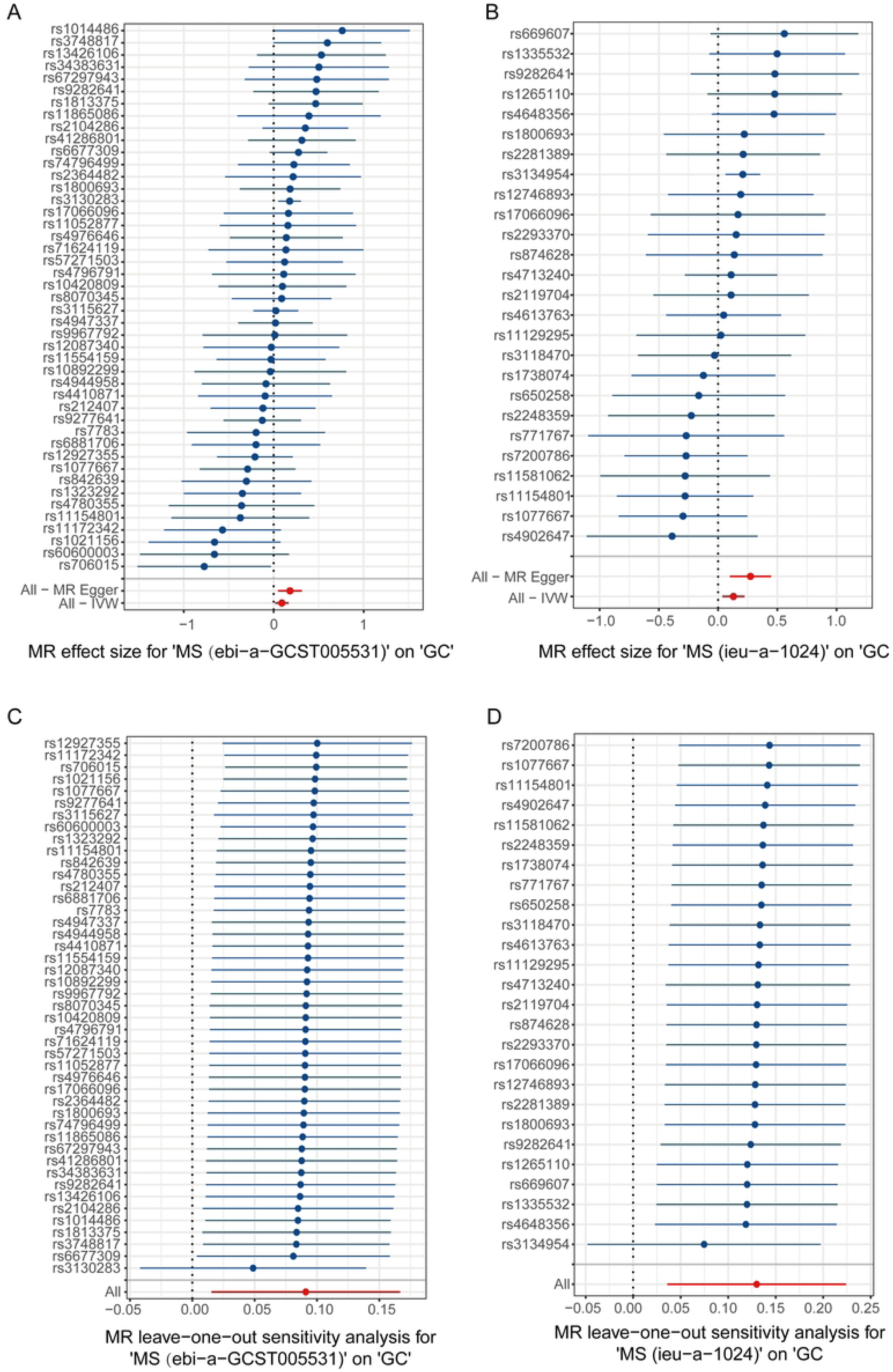
The forest plots show the results (**A-B**) of estimated causal effect for each single nucleotide polymorphism (SNP), and the results (**C-D**) of leave-one-out analysis between multiple sclerosis (MS) and gastric cancer (GC). For A-B, the blue line corresponding to each SNP (begin with rs) represents the causal effect on the outcome in the current analysis direction, where the blue dots represent the value of the causal effect. All and the corresponding red lines represent the overall causal effect in the current analysis direction, where the red dots represent the value of the overall causal effect estimated by different method. For C-D, the blue line corresponding to SNP (begin with rs) represents the causal effect of the remaining SNPs on the outcome after removing the current SNP in the analysis direction, where the blue dots represent the value of the causal effect. All and the corresponding red lines represent the overall causal effect in the current analysis direction, where the red dots represent the value of the overall causal effect. IVW, inverse-variance weighted.

**Table 1.** The results of pleiotropy and heterogeneity test in the Mendelian randomization (MR) analysis between multiple sclerosis (MS) and gastric cancer (GC)

| Exposure | Outcome | Pleiotropy |  |  | Heterogeneity |  |  |
| --- | --- | --- | --- | --- | --- | --- | --- |
|  |  | egger_intercept | SE | P-value | Q test | Q_df | P-value |
| MS (ebi-a-GCST005531) to GC | GC | -0.02 | 0.01 | 0.12 | 47.59 | 44 | 0.33 |
| MS (ieu-a-1024) to GC | GC | -0.03 | 0.02 | 0.07 | 22.12 | 25 | 0.63 |
| GC to MS (ieu-a-1024) | MS | -0.008 | 0.05 | 0.88 | 2.21 | 4 | 0.69 |
| GC to MS (ebi-a-GCST005531) | MS | na | na | na | na | na | na |

na, the results of heterogeneity and pleiotropy analysis cannot be provided due to the acquisition of one SNP in this direction. SE, standard error.

The scatter plots from the forward MR analysis (Fig. 3A-B) demonstrated the causal effects of single nucleotide polymorphisms (SNPs) on GC (Y-axis) and MS (X-axis), along with the overall linear trend that these SNPs fit. The results indicated a significant positive correlation between the exposure and outcome variables. In these analyses, funnel plots (Fig. 3C-D) were used to display the distribution of outliers, showing no apparent outlier values. The scatter plots from the reverse analysis showed the magnitude of causal effects of SNPs on MS (Y-axis) and GC (X-axis), with no apparent correlation observed (Fig. S2A); similarly, the corresponding funnel plot also did not display any potential outlier distribution (Fig. S2B). These results provide a multi-faceted perspective on the causal relationship between MS and GC, evaluating the collective effect of multiple SNPs through different MR methods.

**Fig. 3:**
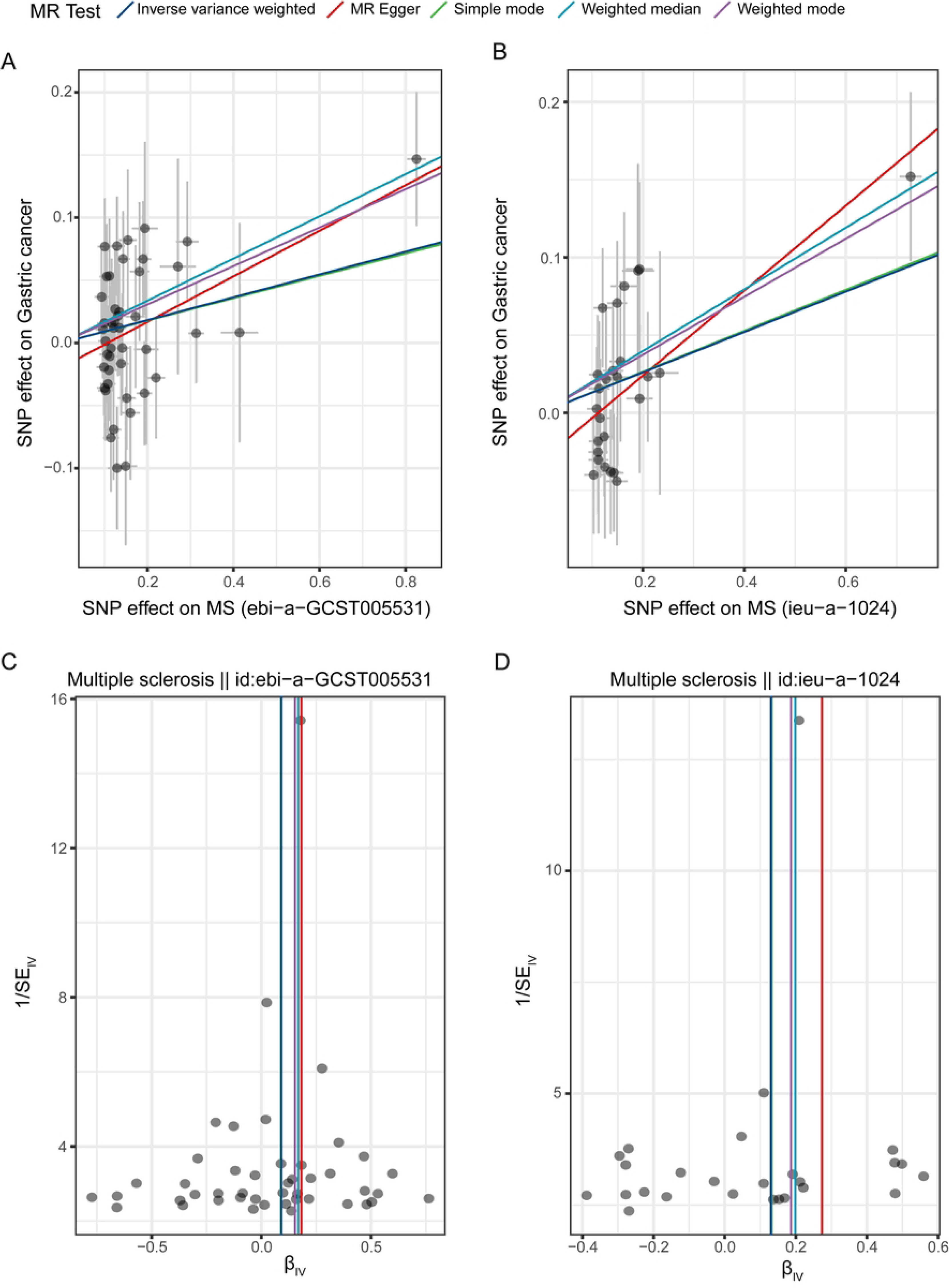
The scatter plots (**A-B**) and funnel plots (**C-D**) show the results of sensitivity analysis in the analysis directions between multiple sclerosis (MS) and gastric cancer (GC). Different lines represent different Mendelian randomization analysis methods, as shown

### DEGs in patients with MS or GC

We retrieved and downloaded sequencing data related to patients with MS and GC from the GEO database, including normal tissue samples matched for each disease as controls. Differential expression analysis conducted on samples from both diseases revealed significant gene expression differences in patients with both MS and GC (Fig. 4A to D). In MS patients, a total of 1,338 DEGs were identified, with 455 genes upregulated and 883 genes downregulated (Fig. 4A, 4B). In GC patients, 1,455 DEGs were identified, with 678 genes upregulated and 777 genes downregulated (Fig. 4C, 4D). The differential genes suggest that they may play a central role in the pathological processes of GC and MS. Specifically, these genes, by regulating the behavior of immune cells, changes in stromal cells, and the release of cytokines, could be key factors in the progression of these diseases and may also represent the biological basis for the interconnection between the two diseases.

**Fig. 4:**
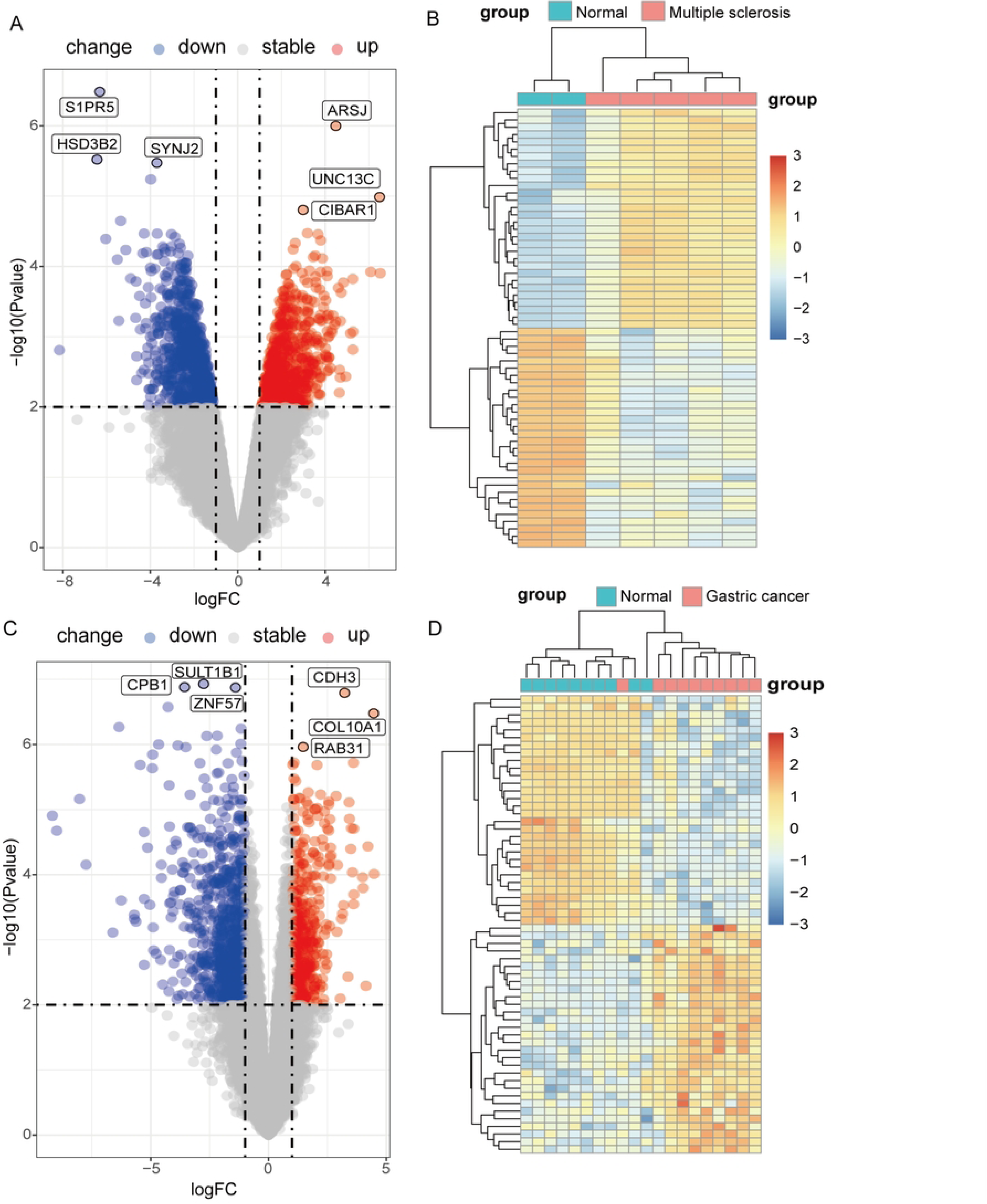
Volcano maps (**A, C**) and heatmaps (**B, D**) respectively display differential genes from patients with multiple sclerosis (MS) and gastric cancer (GC). A-B is for MS, C-D is for GC. The red dots in A and C represent genes that are upregulated compared to their respective normal tissues in two groups of diseases, the blue dots represent genes that are downregulated compared to their respective normal tissues in both groups of diseases, and the gray dots represent genes that are not statistically significant compared to their respective normal tissues in both groups of diseases. Each row in Figures B and D represents a gene, with the redder the corresponding box indicating that the gene is upregulated compared to normal tissues in both disease groups, and the bluer the box indicating that the gene is downregulated compared to normal tissues in both disease groups.

### Enrichment analysis and PPI network of Co-DEGs

In our analysis of MS and GC, we identified 129 co-expressed differential genes (Fig. 5A). GO analysis revealed that these shared differential genes are primarily enriched in pathways such as positive regulation of nervous system development, myelination, and ensheathment of neurons (Fig. 5B). Meanwhile, KEGG enrichment analysis showed significant enrichment in pathways such as Histidine metabolism, Cytoskeleton in muscle cells, and Aldosterone-regulated sodium reabsorption (Fig. 5C). These shared differential genes were uploaded to STRING for PPI analysis and subsequently imported into Cytoscape for network analysis using degree as a measure. Molecular interaction analysis highlighted COL5A2 and THBS2 at the core of the interaction network (Fig. 5D).

**Fig. 5:**
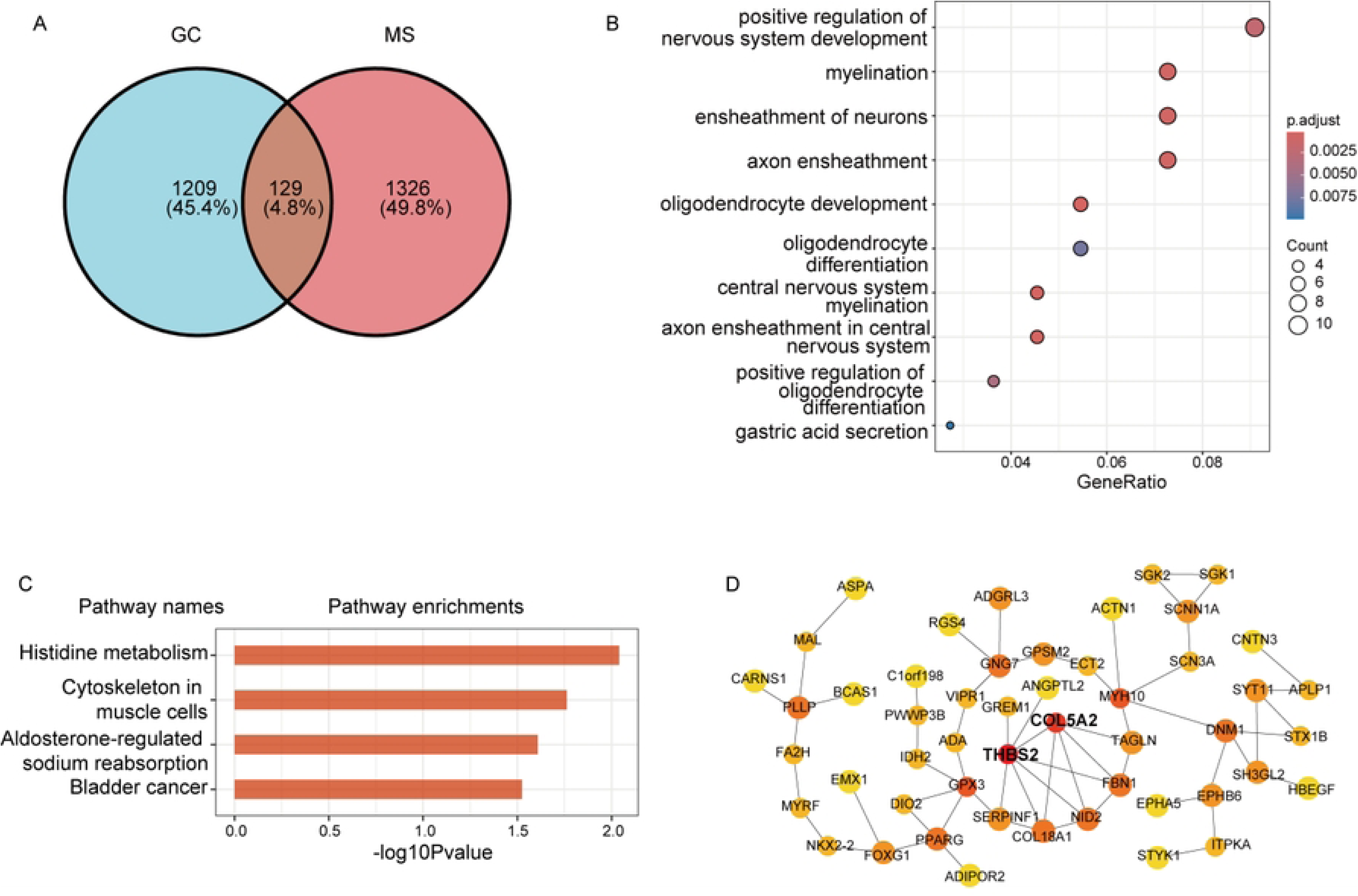
Distribution and enrichment analysis of shared differentially expressed genes (DEGs) in patients with multiple sclerosis (MS) and gastric cancer (GC).(**A**), the Venn diagram shows the number of shared DEGs between these two diseases. (**B**), GO analysis of shared DEGs—biological processes. (**C**), KEGG analysis of shared DEGs—signaling pathways. (**D**), the interaction network of shared DEGs and key genes.

## Discussion

In this study, utilizing MR analysis and transcriptomics data, the results of MR analysis demonstrated a significant positive causal effect of MS on the incidence of GC; however, reverse MR analysis did not reveal a causal effect of GC on MS. These findings reinforce the notion that MS, as a central nervous system disease, may influence the development of GC through its chronic inflammatory processes. Additionally, transcriptomic data analysis identified DEGs shared by MS and GC, particularly those related to immune regulation, stromal formation, and cell migration, further revealing potential molecular links between the two diseases. These results underscore the complex interactions between chronic inflammatory diseases of the nervous system and cancers of the digestive system.

MR analysis is a method that uses genetic variations as IVs to assess the causal relationships between exposures and outcomes^【52】^. In this study, MR analysis confirmed the positive causal effect of MS on increasing the likelihood of GC. This finding is clinically significant, suggesting that individuals with MS should be particularly vigilant about monitoring and preventing GC. From a clinical perspective, as an autoimmune disease, the chronic inflammatory state and the impact on autoimmune factors in MS may be key to influencing the development of GC. For instance, previous studies have reported significantly elevated levels of inflammatory factors such as tumor necrosis factor-alpha (TNF-α) ^【53】^and interleukin-6 (IL-6) in MS patients^【54】^. These factors not only play a key role in the pathology of MS but are also closely associated with the development of GC^【55】^. Elevated levels of inflammatory factors may induce damage and dysfunction in gastric mucosal cells, leading to pathological changes in the gastric mucosa, thereby increasing the risk of GC^【56】^. Additionally, immune system dysfunction in MS patients may lead to decreased immune tolerance to gastric cells, which could also be a significant factor in the development of GC^【57】^.

Moreover, this study employed a variety of bioinformatics tools and algorithms to ensure the accuracy and reliability of results. Statistical methods were used to validate the significance of the identified differential genes and pathways. GO analysis highlighted the main biological processes involved, which encompass a range of key cellular functions such as endoplasmic reticulum calcium transport and negative regulation of axonogenesis, all playing crucial roles in cell development and functional maintenance**^Error! Reference source not found.^**. Furthermore, the regulation of sister chromatid separation and negative regulation of neurite development further indicate the fine control of cell cycle ^【58】^ and neural development ^【60】^that may have cross-impacts in both MS and GC. KEGG pathway analysis further confirmed the involvement of these genes in multiple key signaling pathways, such as Histidine metabolism, Cytoskeleton in muscle cells, and Aldosterone-regulated sodium reabsorption, which are critical in regulating cellular metabolic balance, structural integrity, and electrolyte balance. Particularly in MS and GC, these pathways may promote disease development and progression by affecting inflammatory responses and immune function regulation. For example, the histidine metabolism signaling pathway plays a role in regulating cellular pH balance and intracellular signaling, potentially exerting cross-regulatory effects in the chronic inflammation of MS and oxidative stress responses in GC ^【61】^. Additionally, the involvement of the cytoskeleton signaling pathway in muscle cells may explain the common pathological features of these two diseases in terms of cell structure and dynamics^【62】^.

Through analysis of the differentially expressed and core genes in the dataset within the PPI network, it was revealed that specific key genes (e.g., COL5A2 and THBS2) may serve as molecular links connecting MS and GC. COL5A2, primarily involved in the composition of the extracellular matrix and signal transduction^【63】^, and THBS2, a critical extracellular matrix protein influencing cell proliferation and repair processes^【64】^, may regulate the stability of the cytoskeleton in MS, affecting neuronal cell function. In GC, COL5A2 may influence the adhesion and migration of gastric mucosal cells, affecting gastric mucosal lesions and thereby increasing the risk of GC^【65】^. THBS2 plays roles in regulating cell adhesion and migration and may exacerbate cancer progression in GC by promoting tumor cell invasiveness and angiogenesis^【66】^. These comprehensive analyses not only deepen our understanding of the biological mechanisms of these diseases but also provide potential therapeutic targets, opening new pathways for future clinical interventions and treatment strategies.

Although this study innovatively revealed the causal relationship through which MS increases the risk of GC and delved into the common molecular mechanisms between the two diseases by identifying key genes such as COL5A2 and THBS2, it also presented a precise methodology to reduce the confounding effects commonly seen in observational studies. However, the study has some limitations. Firstly, the results of the MR analysis are highly dependent on the validity and independence of the selected IVs, which need further verification in future studies. Secondly, although transcriptomic data analysis revealed shared differential genes and related biological pathways, these results need to be validated in a broader population and larger sample sizes. Additionally, this study did not fully explore the potential impacts of shared environmental and lifestyle factors between MS and GC, which should be considered in future research.

## Conclusion

Bidirectional MR analysis indicated that Multiple Sclerosis significantly increases the risk of Gastric Cancer, while GC did not show a direct causal effect on the incidence of MS. Moreover, integrative bioinformatics analysis identified key genes, COL5A2 and THBS2, that may play crucial roles in the pathological processes of MS and GC, and are promising as potential targets for future clinical interventions and treatment strategies. These findings not only provide important biomarkers and mechanistic insights into how MS, as a central nervous system inflammatory disease, may influence gastric health through biological pathways but also deepen our understanding of the complex interactions between MS and GC.

## Data Availability

All relevant data are within the manuscript and its Supporting Information files.

## Ethics approval and consent to participate

We employed publicly available GWAS summary statistics data obtained from two databases: the OpenGWAS database (https://gwas.mrcieu.ac.uk/, accessed on May 17, 2024) and the FinnGen database (https://r10.risteys.finngen.fi/endpoints/c3_胃_exallc, accessed on May 17, 2024). Both databases aggregate data from primary studies that have obtained appropriate informed consent from participants and received approval from relevant institutional review boards (IRBs). Since we only used de-identified summary data without accessing individual-level identifiable information, our study does not require additional informed consent from participants.

Ethics statement: Not applicable (relied on IRB approvals and informed consent procedures of the original databases).

Clinical trial number: Not applicable.

## Consent for publication

Not applicable.

## Availability of data and materials

The datasets generated and/or analysed during the current study are available in the ieu open GWAS project (https://gwas.mrcieu.ac.uk/).

## Author Contributions

Conception and design of study: Zhang Z, Wang X, Liao H, Zhang Y, Liao Y

Investigation: Zhang Z, Liao Y

Data analysis and/or interpretation: Liao H, Wang X, Zhang Y

Methodology: Zhang Z, Liao Y

Validation: Liao H, Wang X, Zhang Y

Visualization: Liao H, Wang X, Zhang Y

Formal Analysis: Zhang Z, Liao Y

Supervision: Liao Y

Writing – Original Draft: Zhang Z

Writing – Review & Editing: All authors

All authors read and approved the final manuscript.

## Acknowledgments

We would like to extend our thanks to the OpenGWAS, UK Biobank database, and FinnGen database teams for granting public access to their summary data. Furthermore, we are grateful to the principal investigators of the studies for their transparency in sharing their data for research purposes.

## Supplementary Materials

Figure S1 and S2; Table 1; Tables S1 to S4

